# Text mining biomedical literature to identify extremely unbalanced data for digital epidemiology and systematic reviews: dataset and methods for a SARS-CoV-2 genomic epidemiology study

**DOI:** 10.1101/2023.07.29.23293370

**Authors:** Davy Weissenbacher, Karen O’Connor, Ari Klein, Su Golder, Ivan Flores, Amir Elyaderani, Matthew Scotch, Graciela Gonzalez-Hernandez

## Abstract

There are many studies that require researchers to extract specific information from the published literature, such as details about sequence records or about a randomized control trial. While manual extraction is cost efficient for small studies, larger studies such as systematic reviews are much more costly and time-consuming. To avoid exhaustive manual searches and extraction, and their related cost and effort, natural language processing (NLP) methods can be tailored for the more subtle extraction and decision tasks that typically only humans have performed. The need for such studies that use the published literature as a data source became even more evident as the COVID-19 pandemic raged through the world and millions of sequenced samples were deposited in public repositories such as GISAID and GenBank, promising large genomic epidemiology studies, but more often than not lacked many important details that prevented large-scale studies. Thus, granular geographic location or the most basic patient-relevant data such as demographic information, or clinical outcomes were not noted in the sequence record. However, some of these data was indeed published, but in the text, tables, or supplementary material of a corresponding published article. We present here methods to identify relevant journal articles that report having produced and made available in GenBank or GISAID, new SARS-CoV-2 sequences, as those that initially produced and made available the sequences are the most likely articles to include the high-level details about the patients from whom the sequences were obtained. Human annotators validated the approach, creating a gold standard set for training and validation of a machine learning classifier. Identifying these articles is a crucial step to enable future automated informatics pipelines that will apply Machine Learning and Natural Language Processing to identify patient characteristics such as co-morbidities, outcomes, age, gender, and race, enriching SARS-CoV-2 sequence databases with actionable information for defining large genomic epidemiology studies. Thus, enriched patient metadata can enable secondary data analysis, at scale, to uncover associations between the viral genome (including variants of concern and their sublineages), transmission risk, and health outcomes. However, for such enrichment to happen, the right papers need to be found and very detailed data needs to be extracted from them. Further, finding the very specific articles needed for inclusion is a task that also facilitates scoping and systematic reviews, greatly reducing the time needed for full-text analysis and extraction.

## 1. Introduction

With the massive size of the existing scientific literature and its rapid growth over time, retrieving all pertinent articles for a particular study can be challenging. Researchers dedicate a lot of time and effort to screening titles and abstracts to include or exclude relevant articles and then even more time to extract the pertinent data from the full articles. This whole process is often done manually, with interfaces that facilitate the listing and vetting of the articles, but with little additional automation. Recent advances in Natural Language Processing (NLP) promise to speed up and (semi-)automate this process.

In a recent mapping systematic review, Jimenez et al.^1^ list all tools available to help researchers when writing systematic reviews. Their study concludes that, whereas supporting systematic reviews of the literature with NLP is a dynamic and promising field, technical limitations slow the adoption of the tools by the community. Few tools are available that can be used by researchers that lack a background in Computer Science or programming skills. To be widely adopted, tools need to offer well-designed web or standalone interfaces where most actions can be completed with few clicks and drag-and-drop operations. Such tools are costly to develop, maintain, and update. Web and standalone interfaces require a long time of development and onerous infrastructures to be deployed as well as skilled personnel to maintain the applications and the data generated. Moreover, NLP methods have improved very fast over the last decades thanks to the availability of very large amounts of data from the Internet, more powerful hardware, and better learning algorithms, moving from the earlier rule-based approaches to the current generative approaches powered by large pre-trained neural networks modeling complex linguistic and semantic knowledge. However, the tools available to a general audience are unlikely to integrate the latest technology, since teams managing these tools would more often opt for stability over performance in order to ensure a suitable experience for the hundred of reviewers using the tools during their daily work. For example, the three most cited tools in the list of Jimenez et al.^1^ rely on simple feature-based discriminative classifiers, an approach known to underperform when compared with more recent pre-trained transformer-based neural network classifiers. *Abtsrackr* ^2^ offers a web interface for reviewers to upload a collection of records and screen the titles and abstracts. The tool includes a classifier trained with Active Learning and Dual Supervision algorithms to improve its selection of relevant articles. The classifier discriminates the abstracts based on the predictions of a Support Vector Machine representing the textual content of the articles as N-Grams. *EPPI-Reviewer* ^3^ is a web application integrating multiple tools to facilitate the screening of a collection of articles as well as their analysis. It also integrates a classifier that presents the abstracts to reviewers ranked by relevance given the predictions made by a Support Vector Machine representing the text with tri-Grams. *Rayyan*^4^ is a cloud application helping with semi-automation the screening step of systematic reviews of randomized controlled trials. Like the two previous tools, *Rayyan* predicts relevant articles using a Support Vector Machine representing the abstracts and titles of the articles with stemmed bi and tri-grams along with the MeSH terms discovered automatically.

In this study, we present a general pipeline to screen and extract data semi-automatically from full-texts articles in scientific literature. Our pipeline benefits from recent information extraction techniques to target relevant sentences in full-text articles. This allows us to define detailed selection criteria that are difficult to satisfy, since the relevant piece of information may not be the focus of the published article, but only discussed briefly in one section of the article. We also propose to integrate relation extraction techniques along with active learning to improve the data extraction step. To achieve acceptable performances relation extractors require to be fined-tune on a large number of training examples which, for complex relations, can be very difficult to label. By supporting Active Learning, an interactive and iterative learning method, our extractor will only request the labels of examples of the relations of interest that are the most informative for training the extractor to our human annotators, reducing the workload without compromising the quality of its training. We evaluated the modules of our pipeline implemented on a specific use case: retrieving the metadata for patients infected by SARS-CoV-2 described in the LitCovid collection and for whom the viruses were sequenced and deposited in public databases.

In response to the COVID-19 pandemic, scientists have published over three hundred thousand research articles and made available over 14.6M virus genome sequences.^5^ These sequences, along with their metadata, can be used to understand virus evolution and spread and their implications for public health, a field of study called genomic epidemiology.^6^ For these studies, scientists can leverage databases such as GISAID^5^ and NCBI GenBank^7^ (via NCBI Virus^8^), to access and publish SARS-CoV-2 sequence data. However, SARS-CoV-2 sequence records do not typically contain patient metadata such as demographics, clinical severity, or comorbidities. This prevents researchers from uncovering trends in population health since sequence data across multiple studies that account for a variety of patient cohorts (age, race, gender, comorbidity) and viral genotypes cannot be easily assembled. The sequences metadata are often found in full-text articles. Papers that describe the generation of new SARS-CoV-2 sequences might also describe the characteristics of the patient cohort, their symptoms, and the disease outcomes. Journals require authors to reference the accession numbers of new sequences produced. However, the sequence records in GISAID and GenBank are often missing the reference to the manuscript. Per our preliminary analysis, the vast majority of records are missing a manuscript reference, or only include a reference to a preprint, despite a peer-reviewed paper becoming available after the sequence is published. This might in part be an artifact of how the data is produced: scientists that sequence viruses will generally publish their work, and thus assume that the sequences will primarily be accessed when someone reads the paper. However, in reality, the search for sequence data in genomic epidemiology for secondary analysis starts with the sequence database such as GISAID or GenBank. When the manuscript is found, the metadata, if present, is usually interspersed throughout the text, tables, and supplemental material, and researchers are forced to gather it manually, in a process akin to what we depict in Figure 1. We designed our pipeline to semi-automate the extraction of these metadata.

**Fig. 1:**
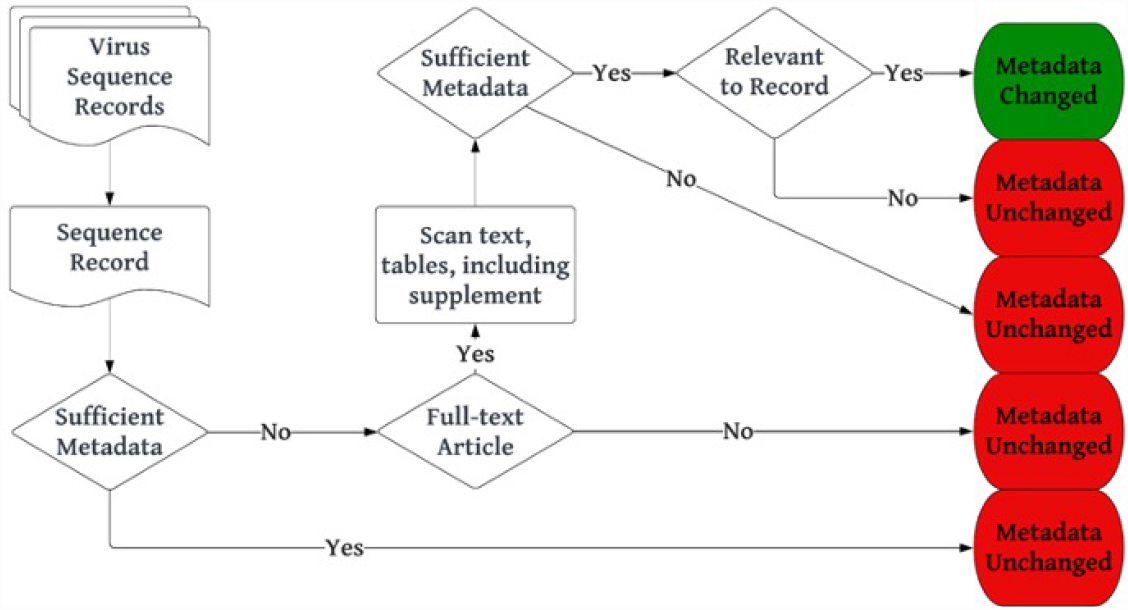
Process to enrich sequence metadata from its corresponding publication.

## 2. Method

As a first step towards linking patient metadata within a journal article to its matching virus sequence record, we developed and validated a semi-automated pipeline to identify relevant journal articles where authors produced and made available in GenBank or GISAID, new SARS-CoV-2 sequences. Figure 2 gives an overview of our pipeline. It is composed of three modules that are applied sequentially to a large collection of articles to retrieve the articles relevant to our study in this collection. Annotators intervene at different steps, either to define the rules to select the relevant articles by defining the linguistic patterns directly with regular expressions, or annotating examples to train a classifier, or to curate the predictions of the classifier on all articles retrieved. In this section, we present the data collection and its annotation, as well as each module of the pipeline.

**Fig. 2:**
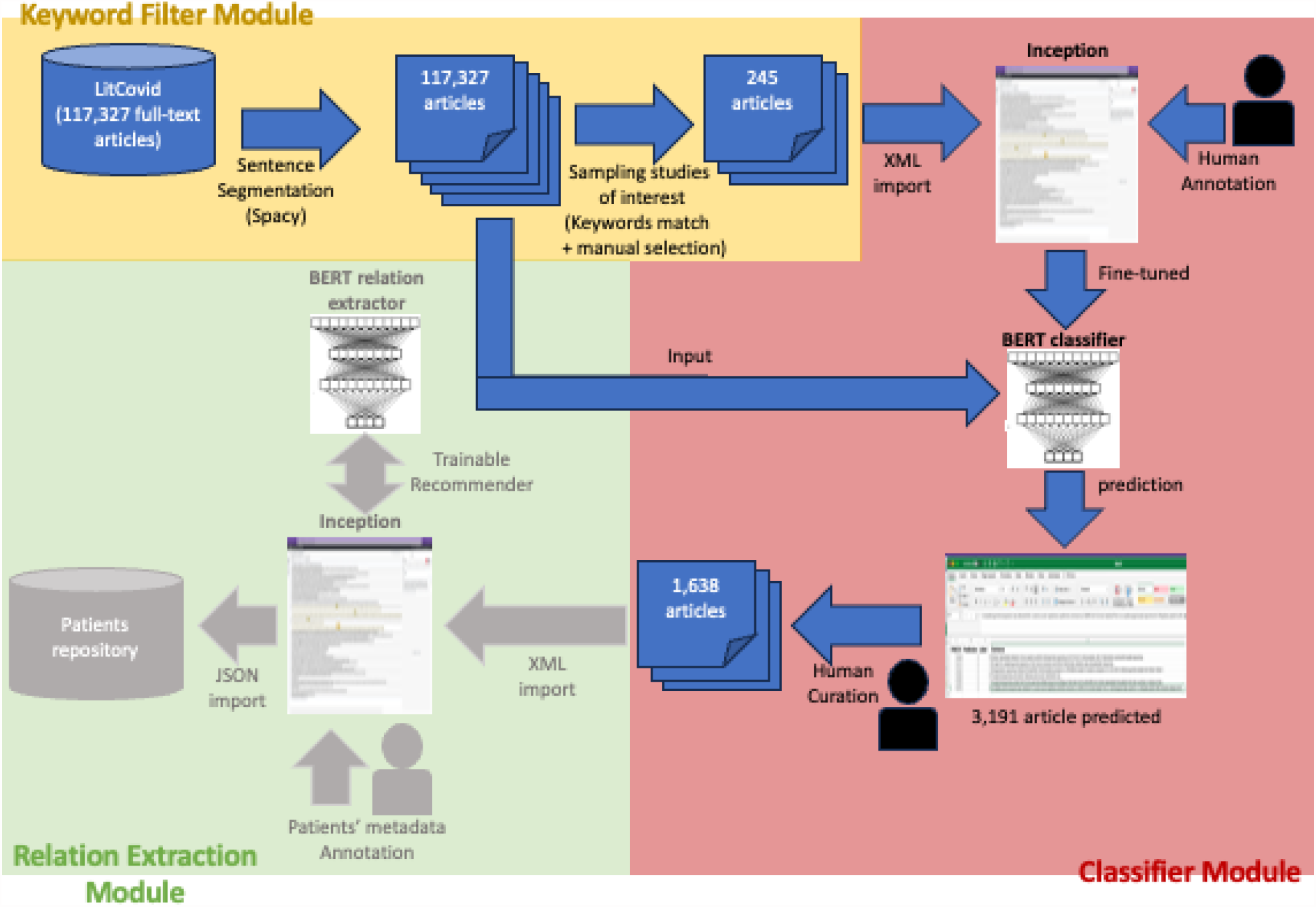
Semi-automatic extraction of SARS-CoV-2 sequence metadata with demographics and clinical phenotypes of patients infected. (*Completed modules are represented in dark blue color, the last module is in progress and represented in grey color*.)

### 2.1 Data collection

An explosion of scientific literature studying the disease shortly followed after the beginning of the SARS-19 outbreak in 2020. In February 2020, the National Library of Medicine (NLM) released a literature hub, called LitCovid, to help healthcare professionals and researchers find relevant scientific information in the mass of publications.^9^ Using a combination of automated and human-in-the-loop methods the authors collected a curated collection of scientific articles related to Covid-19, which at the time of our writing, contained 359,630 articles indexed in PubMed and which is still growing. We downloaded the LitCovid collection through their official API, removing all articles with no full-texts available leading to a collection of 117,327 articles. We preprocessed all articles from this collection to facilitate manual and automatic annotation. The NLM structured the different sections of the articles in the LitCovid collection using tags encoded with the BioC format.^10^ We decoded the structure of the XML articles using the Python library BioC and segmented the sentences of the paragraphs with the Spacy library^a^.

### 2.2 Keyword filter module

We applied sequential filters based on keywords and regular expressions on the NCBI LitCovid collection to identify articles that more likely produced new SARS-CoV-2 sequences. We first excluded papers that did not have a References section to avoid abstract-only entries. Two domain experts reviewed 100 articles to identify keywords and to manually craft regular expressions to be used in the subsequent filtering steps. We identified keywords (Table 1) including laboratory methods (e.g. *synthesized*) and bioinformatics methods (e.g. *MiSeq*), as well as references to the genomic databases GISAID and GenBank. We evaluated our automated approach by randomly selecting 100 articles from the final set (after Filter #4), as well as 100 papers that passed Filter #3 but not Filter #4, to measure recall of the latter. The reviewers noted whether 1) the authors had generated new SARS-CoV-2 sequences as a product of the work, and if so, 2) whether they had deposited the sequences in a public database (GISAID or GenBank), and 3) the estimated number of new virus sequences they produced from the study. The keywords and regular expressions were revised to increase performance. After deploying the final set of keywords and regular expressions, a random sample of 100 articles was reviewed to evaluate the filters, noting the same information as in the prior round.

**Table 1:**
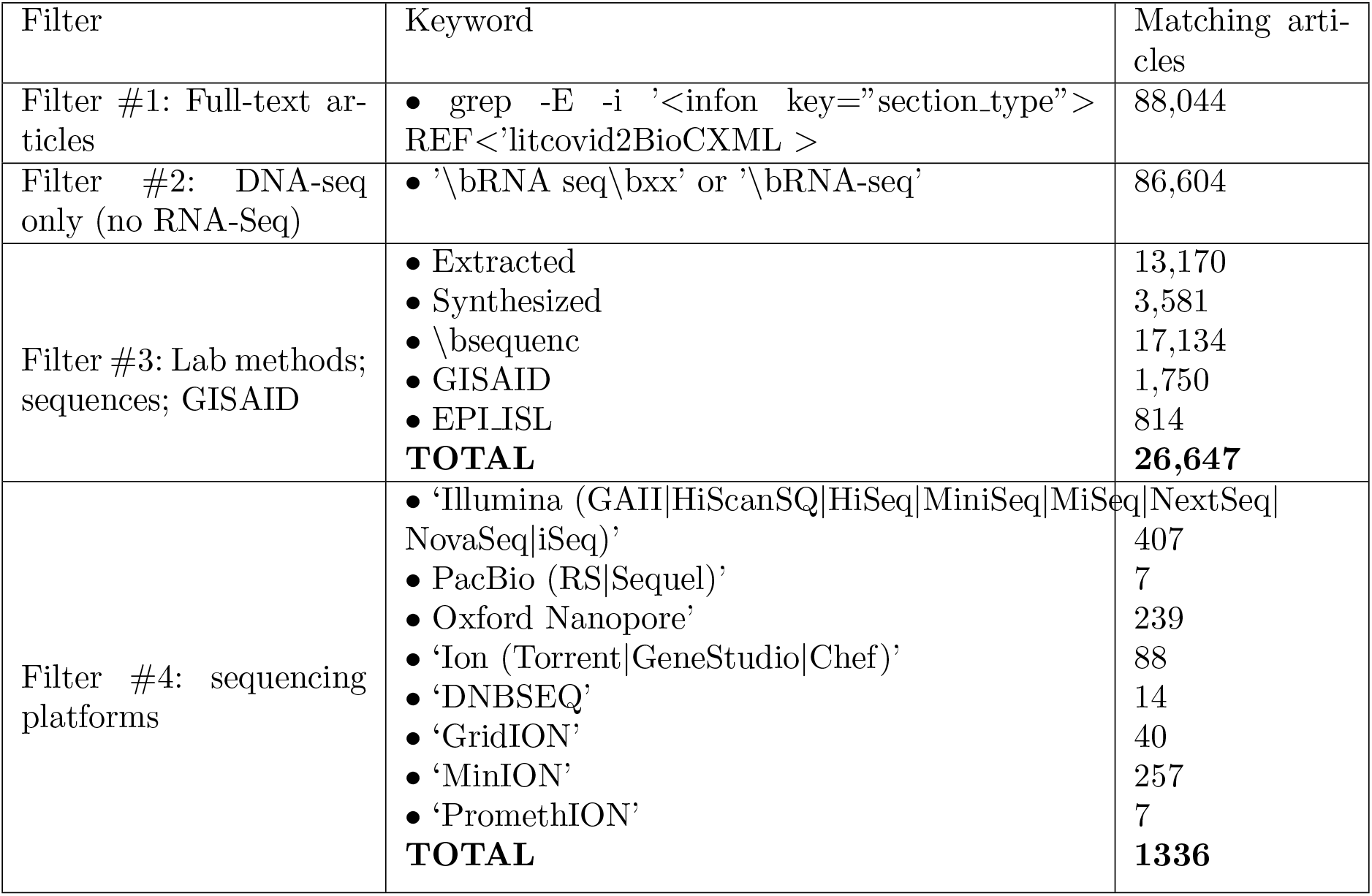
Filters utilized to find articles that report depositing a SARS-CoV-2 sequence in a public repository, with article counts per keyword and TOTAL per filter. Every bullet point keyword reflects an independent count of matching articles at the keyword level. Articles may match several keywords. At the time of our experiments, the LitCovid collection contained 210,059 articles.

We applied our Keyword filter module to the collection of 117,327 articles and randomly selected 245 articles that potentially mention the sequence of a genome. We re-encoded in XML the structures of the articles and imported them into the annotation tool INCEpTION^11^ to facilitate their manual annotation. Our initial annotation task was labeling the articles as relevant, i.e. reporting the generation of new SARS-CoV-2 viral sequences and in accordance with other restrictions in our guidelines (e.g. not sequenced from an animal or wastewater specimen). For articles identified as relevant, annotators selected all sentences of the articles mentioning the sequence of the virus genome (’evidence sentences’) that guided their decision. We conducted several rounds of manual annotations using INCEpTION to train the annotators, measure Inter-rater reliability (IRR), and revise the annotation guidelines as needed. All documents were annotated by two annotators with backgrounds in biomedical informatics. During the development of guidelines, two moderators with domain expertise actively reviewed the annotators’ progress and provided supervision. Once the annotation guidelines were finalized, Annotators 1 and 2 were assigned a random set of identical articles from the curated keyword pipeline findings to establish our Inter-rater reliability (IRR). We measured the classification agreement between them.^12^ We calculated this based on the intersection of one annotator to the annotator with the least number of sentences selected to not penalize agreement for unchosen evidence sentence annotations. All disagreements were resolved by a third annotator with domain expertise.

Through the rounds of training and testing the guidelines, the annotators had near-perfect to perfect agreement (Cohen’s K = 0.95 - 1) in the classification of papers as relevant. For the selection of evidentiary sentences, the agreement improved from arg(R1||R2) = 0.61 to arg(R1||R2) = 0.70 (0.57 - 0.60 Cohen’s kappa). After adjudication and discussion, the guidelines were deemed finalized and the annotators were given a set of 75 articles to assess agreement on the finalized guidelines. The annotator agreement for the classification of the 75 articles was K = 1.0, indicating perfect agreement. The agreement of the evidence sentences selected among these 75 articles was arg(R1||R2) = 0.72 (Cohen’s kappa = 0.71) indicating moderate agreement.^13^

Note that the keyword matching method is commonly used to retrieve relevant articles for a study and it is often the only method applied for the search. We also used keyword matching in our study however, but due to the limitation of this method, we did not use it as a retrieving method but as a sampling method. The goal of the first module was only to help us select a small set of relevant articles for annotation to train a classifier, the main component of the second module. In fact, the keyword matching method only retrieves the articles which have a sentence containing a mention of a keyword from a manually pre-selected list of keywords. But, because any list of keywords is unlikely to be complete for complex queries such as our query, this method misses articles relevant to our study, resulting in many relevant articles missed (False Negative). Moreover, this method searched for the keywords in isolation from the sentences or paragraphs where they appear, making it impossible to distinguish articles where the keywords take a meaning different from the meaning intended when we added the keywords to the list, adding many irrelevant articles (False Positive) to the articles retrieved. We reduced both False Negative and False Positive articles by improving the search method using Machine Learning in our second module.

### 2.3 Classifier module

The main component of the second module, the classifier module, is a classifier trained to detect the sentences reporting viral genome sequencing and to ignore the other sentences. We split the 245 annotated articles selected by the first module into three sets, a training set that contains 60% of the articles (147 articles, 31,885 sentences), a validation set with 20% (49 articles, 9,017 sentences), and a test set with 20% (49 articles, 10,017 sentences). Our corpus is challenging for training a classifier since it is extremely imbalanced^14^ with only 357 sentences reporting viral genome sequencing, the positive examples, and 50,561 which did not, the negative examples. Extremely imbalanced corpus, where training examples belonging to one class outnumber the training examples from other classes, are known to be challenging for standard learning algorithms which assume an equal distribution between the examples. The first challenge caused by the imbalance is that the cost of annotation increases. Among the 245 articles, and despite our pre-selection with the keywords from our list, we found that only 74 articles were mentioning a sequencing process of a viral genome. Our annotators spent most of their efforts annotating articles with no positive examples needed to provide information about the linguistic patterns used by researchers to report viral genome sequencing. The second challenge caused by the imbalance is algorithmic. The standard learning algorithms designed to learn from balanced datasets tend to overgeneralize when applied to imbalanced datasets. They classify the few positive examples of the training set as negative examples in order to minimize their loss during training and achieved poor performance when evaluated on the positive class.

As a baseline, we trained and evaluated a support vector machine (SVM) classifier, using the LibSVM implementation in Weka. We pre-processed the sentences by normalizing URLs and numbers, removing non-alphanumeric characters and stopwords, and lowercasing and stemming^15^ the text. We extracted n-grams (n=1-3) as features in a bag-of-words representation. We used the radial basis function kernel and set the *cost* parameter at *c*=32 and the *weights* parameter at *w* =1 for the “negative” class and *w* =30 for the “positive” class, using the validation set to optimize these parameters. As the main classifier, we instantiated a pretrained neural network from the Hugging Face^b^, a transformer model BERT-base-uncased,^16^ and fine-tuned it to perform the task. We trained the initial BERT model for 20 epochs and reloaded the model which achieved the best F1-score on the validation set, before evaluating its performance on our test set. We evaluate the performance of our classifiers with the standard F1, precision, and recall metrics computed on the positive sentences. To improve the training of our BERT classifier on our extremely imbalanced corpus, we tried undersampling our training set by removing all negative examples in the set which were too dissimilar from the positive examples. We first computed the embeddings of all sentences of the training set with the pre-trained network sentenceTransformer.^17^ Then, we kept in our training set all positive sentences and removed all negative sentences which have their embeddings located far from the embeddings of all positive examples in the n-dimension space of the feature space defined by the embeddings. Finally, we retrained our classifier on this undersampled training set and evaluated the best model on our test set.

Once our BERT classifier was trained and evaluated, we classified the sentences of the 117,327 articles in our collection to retrieve articles most likely to discuss a viral genome sequencing process. We stored the sentences predicted as positive in an Excel file to help the manual curation of our results and ensure that all articles curated were relevant articles. We hypothesized that our classifier would retrieve more articles of interest and with less noise than a simple keyword-matching search, making a manual curation of the predictions possible. Our neural network allows a better representation of the context where the keywords are mentioned, efficiently rejecting negative sentences with keyword occurrences and detecting complex linguistic patterns involving synonyms or hyponyms/hyperonyms not present in our initial list of keywords. We also expected to miss relevant articles with our classifier since we trained it on a limited set of positive examples, a set unlikely to contain all possible ways to refer to a viral genome sequencing process. Therefore, in addition to the recall we measured on our test set, we also estimated the number of relevant articles in our collection missed by our classifier by looking at a particular subset of articles in our collection, the subset of all articles that have a link to a viral genome sequence deposited in GenBank by the authors. These articles are relevant articles that should have been detected by our classifier.

### 2.4 Relation extraction module

We are currently implementing the last module of our pipeline. This module will extract from the relevant articles detected by the Classifier module all concepts of interest describing the patients infected by a SARS-CoV-2 virus for which the genome was sequenced and described in the articles. These concepts include any mentions of the demographics information of the patients (age, sex, race/ethnicity), relevant clinical information (symptoms presented, treatment with medications given and dosage, comorbid conditions, and outcome of the infection which are labeled as asymptomatic, mild, hospitalized, ICU/Ventilator or fatal), as well as geographic metadata when available (location of collection of the virus, travel history of the patients and their places of residence). If the piece of information is present, the module will also link the GISAID or GenBank sequence IDs to the patient whose information was extracted. We define this task as a relation extraction task where an annotator/system should detect the concepts of interest and link them with the appropriate relations. For example, in the sentence ”*A 35-year-old woman, who lives in Wuhan, China, arrived at the Incheon Airport on January 19, 2020*.”, the phrase *woman* should be identified as the patient, *35-year-old* as age, and the relation *has years of age* linking both concepts. The manual annotation of relations can be time-consuming and error-prone. We are currently developing an external recommender for Inception^11^ to, both, reduce this annotation effort and improve the performance of the recommender. Our recommender implements an active learning interface.^18^ Following this training method, we will train our recommender through multiple iterations where the recommender will interact with an annotator. At each iteration, our recommender will search the entire corpus of articles for the paragraphs that may mention the relations but are difficult to predict. These relations are the most informative for its training. The recommender will submit the suspected relations to the annotator through the Inception annotation interface for correction and will retrain its extractor once all relations are corrected. After several iterations, we hypothesize that our recommender will detect most relations and achieve better performance than if it was trained on the same number of relations but randomly chosen.^19^ The relations extracted will be exported into a dedicated database for further genomic epidemiology studies.

## 3. Results

Our baseline SVM classifier achieved an F_1_-score of 0.370 (precision=0.329 and recall=0.422) for the class of sentences that mention the sequencing of a viral genome. Our deep neural network classifier substantially outperformed this baseline, with a 0.480 F1 score, 0.492 Precision, and 0.469 Recall. Unfortunately, our undersampling approach did not improve the performance of our classifier, with a 0.453 F1-score, so we kept the simpler model trained on all examples for our pipeline. Although our classifier achieved moderate performance at the sentence level, it achieved very high performance at the article level in our test set, with 0.80 F1 score, 0.667 Precision, and 1.0 Recall. In the 49 articles composing our test set, our classifier correctly detected all 12 articles of interest and only predicted 6 irrelevant articles as articles of interest. Moreover, in the 12 articles detected, the classifier always identified at least one sentence labeled as positive by our annotators. This was important when we manually curated the predictions made by our classifier on the 117,327 articles. The annotator could only look at the list of sentences predicted positive by the classifier and not the full article to decide if the article was relevant since it was very likely that a sentence mentioning the sequencing process was detected by the classifier and appended to the list. However, the perfect recall of our classifier is most likely due to the pre-selection of the articles composing the test set with keywords which are quickly learned by the classifier. We calculated a lower recall with our heuristic. To better estimate the recall, we used the e-Utilities of NCBI to find publications linked to a SARS-CoV-2 accession numbers for creating a gold standard to compare our classifier against. From the 7,8 million sequences accessed using the SARS-CoV-2 datahub, 9049 sequences were linked to 133 unique PubMed articles, with 58 articles included in the LitCovid collection run through our classifier. Of those 58, our classifier identified 28 correctly (True Positives) and improperly classified 7 articles of interest as negative (False Negatives’), achieving a lower, yet, still high 0.800 Recall.

Within our initial collection of 117,327 articles, our classifier predicted that 3,191 articles were of interest. After manual curation, we found that 1,621 articles were actually relevant, achieving 0.508 Precision. We analyzed the types of errors made by our classifier. We found 10 categories of error types. The largest type (n = 645) was the selection of sentences not related to sequencing based on a word found in sequencing sentences, such as ”obtained” or ‘assemble’. Other errors included the selection of sentences describing or discussing another sequencing effort (n = 245), discussions of retrieved sequences (n = 123), and discussion of testing or positive samples (n = 90).

## 4. Discussion

In this study, we introduced a pipeline designed to search for articles mentioning the sequence of SARS-CoV-2 virus infecting patients. This selection criteria is a minor detail in the articles of interest that focus on larger studies. Therefore, the key terms needed to retrieve these articles are unlikely to be mentioned in the titles or the abstracts which are often the only sections of the articles screened by researchers when following the standard protocol to conduct systematic reviews.^20^ Our pipeline offers an alternative solution to keyword-based searches in titles and abstracts by applying modules implementing current information extraction methods. These modules parse automatically the full text of the articles to retrieve the specific facts in relevant articles. Whereas the first module of our pipeline also implements a keyword-based search, the module only retrieves a sample of articles of interest to enable the annotation needed to train the classifier of the second module which is in charge of the automatic screening of the articles. We were able to retrieve all relevant articles that were selected with keywords in our test set and estimated a good score of 0.80 Recall on the articles not matched by our keywords in the LitCovid collection.

Our pipeline is still imprecise despite the improvement offered by its classifier module. It requires a substantial effort to manually curate its predictions since it retrieves many articles that were not relevant. We found during our analysis of the errors that a large number of False Positives predicted by the classifier were negative sentences similar to positive sentences in the training set. The annotators had to read the entire paragraphs to find the piece of information needed to label these sentences as negative. Our classifier relies on a BERT transformer to represent the meaning of a text. Due to computation limitations, the size of the transformer neural network cannot encode text longer than 512 tokens (i.e. words or segments of words), making the classifier unable to classify more than one long sentence at a time and, without access to the full paragraph, it cannot solve our task which often requires inter-sentence relationship understanding. This strong limitation has been recently lifted with the availability of better hardware and algorithms, leading to the release of very large language models by the community. For example, GPT-4 which powers chatGP,^21,22^ accepts inputs of 8000 tokens and it can be extended to 32k.^23^ We plan, as future work, to encode the entire paragraphs as input for our classifier module by replacing the BERT model with a larger language model in open-access such as BLOOM.^24^

Integrating a large language model, like BLOOM, in our pipeline would have another benefit besides the improvement of its precision. We hypothesize that a large language model will increase the genericity and re-usability of our pipeline. The current pipeline screens the literature using a BERT classifier. This classifier needs to be trained on a large number of examples to achieve acceptable performance. This training limits the re-usability of our pipeline if one wants to change the selection criteria for the need of another study. Annotations still involve human annotators, have a high cost, and once the examples are obtained, retraining the classifier on these examples involves specialized technicians and hardware availability. Instead, the larger size of the layers composing the networks of large language models allows them to encode long-range dependencies between sentences of a paragraph, thus capturing the meaning of the text. When large language models are pre-trained on massive corpora composed of documents from multiple domains, they learn linguistic and semantic knowledge they can recall on demand to perform prediction tasks that they have never been explicitly trained on.^25^ This property makes some authors, such as Moor et al. in,^26^ compare these models with generalist intelligent agents. LLMs can learn in the context of a few examples provided by a researcher to perform new tasks and achieve noticeable performance. We plan to add a user interface in our pipeline to help researchers interact with a large language model and define some examples of relevant and irrelevant articles for their study which should be enough for the model to screen the literature without being explicitly trained to do so.

## 5. Conclusion

We presented a pipeline to identify articles in the scientific literature that report having produced and deposited in GenBank or GISAID new sequences of SARS-CoV-2 viruses infecting human patients, a crucial step to enable our pipeline to extract patient characteristics such as co-morbidities, outcomes, age, gender, and race, and enrich SARS-CoV-2 sequence databases where these pieces of information are often missing. The gold standard set we created during this study is available for training and evaluating new machine learning classifiers. Beyond our particular case study, we showed that, with the integration of an information extraction module in our pipeline, we could retrieve automatically and efficiently articles discussing a minor piece of information within the context of a larger study. We are currently evaluating large language models that have been proven to perform new tasks with few examples, a property that should facilitate the large adoption and utilization of our pipeline.

## Data Availability

All data produced in the present study are available upon reasonable request to the authors

## Acknowledgements

Research reported in this publication was supported by the National Institute of Allergy And Infectious Diseases of the National Institutes of Health under Award Number R01AI164481 to GG and MS

## Footnotes

a Available at https://spacy.io/api

b Available at https://huggingface.co/bert-base-uncased

## Notes

### Competing Interest Statement

The authors have declared no competing interest.

